# Diazinon residues levels in farm-gate *Brassica oleracea* var. *acephala* of Kimira-Oluch Smallholder Farm Improvement Project, Kenya

**DOI:** 10.1101/2024.09.05.24313144

**Authors:** George Odoyo Oromo, Philip Okinda Owuor, Bowa Kwach, Peter Otieno

## Abstract

Pesticides use in vegetable production often has residual effects on the plants and environment with potential health risks. Diazinon, though associated with human health impacts, is a popular pesticide in the production of *Brassica oleracea* var. *acephala* at the Kimira-Oluch Smallholder Farmers Improvement Project (KOSFIP), Kenya. The long preharvest interval (PHI) of diazinon application may not be observed by farmers with inadequate appreciation of Good Agricultural Practices (GAP). It is not documented whether diazinon residues levels in the farm-gate *Brassica oleracea* var. *acephala* of KOSFIP could be a health risk to the consumers. The diazinon residues levels and corresponding health risks in farm-gate *Brassica oleracea var, acephala* at KOSFIP were determined. Cross-sectional survey based on snowball sampling identified 40 farms applying diazinon on the vegetable. Triplicate samples were collected from each farm for residue analysis, using the QuEChERS method, and LC-ESI-MS/MS analysis. Standard normal distribution function f(z), revealed ≈ 78% of farm-gate samples had detectable residual diazinon levels and 70% were above the Codex MRL of 0.05 mg/kg. The farm-gate *Brassica oleracea var. acephala* are exposing consumers to health risks. Efforts must be intensified to ensure GAP are adopted. The estimated farm-gate samples with health risk indices for children and adults (HRIc and HRI_A_) >1.0 were 64% and 26%, respectively. Farm-gate *Brassica oleracea var. acephala* diazinon levels are therefore causing high health risks to both children and adults. Farm-gate residual levels and HRI were comparatively higher than findings of most previous studies. Inappropriate label PHI and malpractices against GAP may be responsible for high residual levels. There should be regular surveillance and trainings of farmers on GAP for sustainable production of *Brassica oleracea* var. *acephala* in the Lake Victoria region. Use of diazinon on *Brassica oleracea* var. *acephala* should be discouraged and alternative approaches including integrated pest management practices should be encouraged.

## INTRODUCTION

Pesticides enhance crop production for the increasing global population towards the realization of sustainable development goals (SDGs) of the United Nations (1,2). However, synthetic pesticides residues, magnified by malpractices in use, are ubiquitous contaminants in the environment (3–5). The residues pose serious to lethal health hazards to non-target organisms through inhalation, contact and ingestion of the contaminated foodstuffs (6,7). Approximately 30% (based on mass) of human food is of vegetable origin, mostly consumed raw or semi-processed. Vegetables can therefore be sources of pesticide residues to human beings more than other food groups (8) since most vegetable production uses various pesticides. Ingestion of contaminated foodstuffs is a major exposure route to pesticide residues (9). It is necessary that vegetables treated with pesticides during production are evaluated for pesticide residue safety levels to protect the consumers against food safety hazards and risks.

*Brassica oleracea* var. *acephala* is grown in many parts of the world (10,11) as food and for its numerous health benefiting metabolites that minimize the risk of degenerative diseases like cancer (6,12). However, cultivation of the vegetable involves application of pesticides for the management of pests and diseases that attack the roots and foliage (13–15). In Kenya, diazinon (*O,O*–diethyl–*O*- (2-isopropyl-6-methyl-4-pyrimidinyl)phosphorothioate) is one of the broad-spectrum pesticides registered for use in *Brassica oleracea* var. *acephala* production (16). Diazinon is expected to undergo dissipation through chemical degradation processes and other forms of physical transformations including surface wash-off from the leaves. The total dissipation effect are expected to reduce diazinon residues to levels below the Codex maximum residue limit (MRL) of 0.05 mg/kg when diazinon is applied according to good agricultural practices (GAP) (17).

Kimira-Oluch Smallholder Farmers Improvement Project (KOSFIP) is an irrigation project located in Homa Bay County in the Republic of Kenya. The area is characterized by hot and humid climatic conditions with scanty rainfall of high variability in duration and amounts (18,19). High relative humidity of the location is due to close proximity to Lake Victoria and the irrigation water channels of the project. These conditions promote rapid spread of vegetable pests and diseases (20,21). The smallholder farmers manage the pests by use of synthetic pesticides. Though diazinon has been used by the smallholder farmers of *Brassica oleracea* var. *acephala* at KOSFIP, the long pre-harvest interval (PHI) of 12 days (16) may not be observed by the farmers (5). Consequently, it is possible that residues of diazinon in *Brassica oleracea* var. *acephala* grown in KOSFIP may be above Codex MRL. Unfortunately, farm and market gate basket screening of diazinon residues in the produce at KOSFIP is not documented. The diazinon residues in the farm gate *Brassica oleracea* var. *acephala* vegetables might be a health risk to the consumers. Consequently, a survey of diazinon residue levels in farm gate vegetables and the health risks the residue levels may pose were investigated. The survey results of farm gate diazinon residues in *Brassica oleracea* var. *acephala* vegetables are reported herein.

## MATERIALS AND METHODS

### Study area

This study was carried out within the Kimira-Oluch Smallholder Farmers Improvement Project (KOSFIP) site. KOSFIP is an irrigation project located in Rachuonyo (Kimira site) and Homa Bay (Oluch site) sub-counties of Homa Bay County in Kenya (Figure 1)(22). The site lies between latitudes 0° 20’ S and 0° 30’ S and longitudes 34° 30’ E and 34° 39’ E at an altitude of 1154 m above mean sea level along the shores of Lake Victoria (Figure 2)(23). Kimira site has an area of 1,790 ha out of which 808 ha have been developed into 44 farming blocks whilst Oluch site has an area of 1,308 ha with only 666 ha split into 53 farming blocks have been irrigated (19). The area is sub-humid with mean annual rainfall of between 740 and 1200 mm with short and long rainy seasons during April-May and November-December, respectively, and mean annual maximum temperatures of 31^0^C and minimum of 18^0^C (18,19). The relative humidity is significantly high ranging between 60 and 75% with potential evapotranspiration rate at 1800mm and 2000 mm per annum. Apart from the rains, the farms are irrigated, thereby producing vegetables throughout the year. The sites have fertile alluvial soils originating from the nutrient alluvial deposits washed downstream from the rivers and erosions from the Gusii highlands.

### Research design

A cross-sectional survey design based on purposive and snowball sampling techniques were used to identify forty-five farms of *Brassica oleracea* var. *acephala* that use diazinon in vegetable production. The survey was carried out during dry season (February to March, 2020) in both Kimira and Oluch sites of the project. During the period, Kimira site had 18 active blocks with *Brassica oleracea* var. *acephala* while Oluch had 35 sites. In addition, during sampling, Kimira site had 12 farms while Oluch had 33 farms. The criteria for snowballing included same vegetable variety treated with diazinon only at first harvest. Among the forty-five farms, 40 were selected (24). Kimira had 11 farms while Oluch had 29 farms. The farms were spread equitably to represent Kimira and Oluch sections of the project, considering all the 97 blocks making the project area. Sampling was biased to farms of *Brassica oleracea* var. *acephala* that had been treated with diazinon before the first harvest after planting. From every farm, 1 Kg of freshly harvested *Brassica oleracea* var. *acephala*, replicated three times were collected to make 120 samples.

### Sample processing, preparation, extraction and partitioning

The processing, preparations, extraction and partitioning of the vegetable samples for diazinon residues analysis were carried out using Quick Easy Cheap Effective Rugged and Safe (QuEChERS) multi-residue method (25) as adopted and validated by the Analytical Chemistry Laboratories of Kenya Plant Health Inspectorate Services (KEPHIS) method M0326 as follows.

### Sample processing and preparation

The vegetable samples were coarsely cut with a knife then chopped and homogenized with a Hobart food processor. About 100 g of the homogenized samples were placed in sample containers and were then stored frozen at −18°C ± 5°C in readiness for extraction.

### Extraction and partitioning of samples

A 10.0 ± 0.1 g of the homogenous wet samples and controls were weighed into 50 ml centrifuge tubes. The control samples were fortified with diazinon standard solution to achieve the 0.01µg/g for LC-MS/MS. Using an automatic pipette, 50 µl of procedural internal standards (dimethoate D6 (10 ppm) and malathion D10 (10 ppm)) were added to the contents of the centrifuge tubes. To the contents of the centrifuge tube, 10.0 ± 0.2 ml extraction solvent acetonitrile (MeCN) HPLC grade was added. The tube was immediately closed and shaken vigorously by Geno grinder for 1 minute at 1000 revolutions per minute (rpm). The resulting homogenous mixture in the centrifuge tube was then subjected to liquid-liquid partitioning step using 6.5 g of premixed extraction salts. The extraction salts comprised 4.0 ± 0.2 g magnesium sulphate anhydrous for removal of water and salting out MeCN; 1.0 ± 0.05 g sodium chloride to increase selectivity of analyte by reducing amount of co-extracted matrix; 1.0 ± 0.05 g trisodium citrate dihydrate and 0.5 ± 0.03 g disodium hydrogen citrate sesquihydrate as a citrate buffer for pH adjustment. The tube was closed and immediately shaken vigorously by hand to avoid caking. The mixture was again shaken by Geno grinder for 1 minute with 1000 rpm then centrifuged for 5 minutes at 3700 rpm. An aliquot of 500 µl of the mixture was transferred into a 2.0 ml vial followed with 495 µl of HPLC grade water and 5 µl of injection internal standard dimethoate D6 (10 ppm). The mixture was vortexed to mix for LC-MS/MS analysis. The extracts were directly subjected to quantitative analysis by LC-MS/MS (dMRM) mode.

### Preparation of calibration solutions

Calibration solutions were prepared using a control matrix containing no detectable residues of diazinon analytes. Using an automatic pipette, 4 ml of control blank was put into a 15 ml centrifuge tube followed with 4 ml of HPLC grade water and vortexed to mix. Reference standard solutions of diazinon pesticide stocked by KEPHIS were prepared for analysis at concentrations of 0.005 µg/ml, 0.01 µg/ml, 0.02 µg/ml, 0.05 µg/ml, 0.075 µg/ml, 0.1 µg/ml and 0.2 µg/ml for validation of method, and 0.005 µg/ml, 0.02 µg/ml, 0.05 µg/ml, and 0.2 µg/ml for routine analysis using the blank control.

### Instrumentation and instrument specifications

The extracted samples of *Brassica oleracea var. acephala* were analysed using Liquid Chromatography Quadruple Agilent 6430 LC-MS/MS (HPLC) with standard electron spray ionization (ESI). The HPLC column used was C-18 with an internal diameter of 1.8 µm and a length of 50 cm. The optimization parameters, including solvent gradient, precursor and product ions, and retention times were set as outlined in method M0326. The column temperature was set at 40.0°C. The auto-sampler injection and ejection speed was 200µL/min with an injection volume of 3.00 µL

### Quality control

The 2018 Standard Operating Principles (SOP) number M0326 for QuEChERS Multi-Residue Method for Analysis of Pesticides Residues (25) in high water matrices validated by Analytical Chemistry Laboratory of KEPHIS was used in the determination of residue levels. The quality parameters included repeatability, linearity, accuracy of recovery, method’s limits of detection (LOD) and quantitation (LOQ). Calibration curves were drawn according to analyte ranges of concentration and response to the LC-ESI-MS/MS. This was achieved by using five replicates of different concentrations diluted with blank extract samples. Evaluations of accuracy and precision parameters were done by recovery experiments (recovery range of 93 – 123%) in which each analyte standard were spiked with blank *Brassica oleracea* var. *acephala* slurry in six replicates. The replicates were prepared separately at three different concentrations of 10, 20 and 50 μg Kg^-1^. Limits of detection (LODs) of each analyte was validated by comparing the signal-to-noise (S/N) ratio magnitude to the background noise obtained from blank sample in the six replicates that presented mean coefficient of variations (CV) of less than 20%. The time window for the signal – to - noise (S/N) ratio was set at t < 2 minutes. LOD was calculated using the mathematical expression (26):

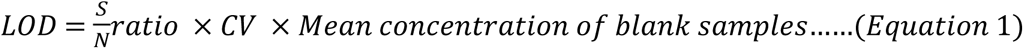

The limits of quantification (LOQs), defined as the minimum concentration of an analyte that can be identified and quantified with 99% confidence, was calculated using the mathematical expression (26):

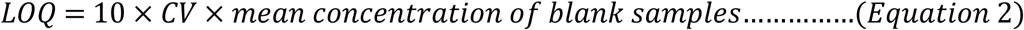

### Analytical determination of residual diazinon

For quantitative analysis of the analytes, 3 µl of the solvent, matrix control, calibration standards and spikes, and samples were injected into the LC-MS/MS instrument. Responses were recorded for both internal standards and samples. A calibration curve of responses against concentration of calibration standards was obtained. The results of concentrations of residues of diazinon for all the samples were calculated from responses obtained from the calibration curve. Respective chromatograms and graphics for quantitation and confirmation were obtained.

### Statistical analysis

The cross-sectional survey data of diazinon residues in farm gate samples were subjected to descriptive statistics for purposes of illustrating measures of central tendency and dispersion: mean concentrations, mode, median, quartiles, standard deviations, minimum and maximum values, range of values and the coefficient of variations (CV). The diazinon residues levels were also compared with Codex MRL values of 0.05 mg/kg and evaluated for health risk assessment. For both residue levels and health risk indices, the standard normal distribution function f(z) was used to determine the proportion of values that lie below and above the tolerable values of MRL and HRI. The values were computed using the formula:

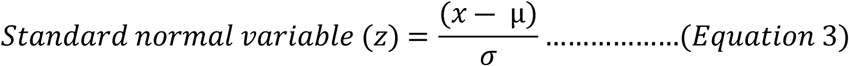

Where z is the standard proportion value; x is the acceptable value; µ is the mean of the data set; and, σ is the standard deviation of the data set.

### Residual Pesticide Health Risk Assessment

The residual pesticide health risk indices (HRI) estimations for children (HRI**_C_**) and for adults (HRI**_A_**) based on the farm gate samples were estimated using the European Union formula (27):

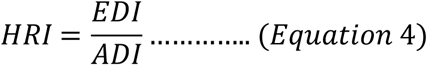

Where; HRI is the health risk index; EDI is the estimated daily intake of the pesticide; and ADI is the acceptable daily intake of the pesticide.

The EU formula provides that the EDI be determined by multiplying the sample residual pesticide concentration (mg/kg) by the estimated WHO food consumption rate (kg/day), and dividing by the number of the estimated WHO average body weight (28). HRI > 1.0 were considered as posing health risks hence not safe for human health. HRI ≤ 1.0 were considered not posing immediate health risks and thus safe for human health (29,30). The average daily vegetable intake for an adult of average weight 60 Kg was considered to be 0.345 Kg/person/day while children average daily intake was considered to be 0.232 kg/person/day for average body weight of 10 kg (31). The maximum acceptable daily intake (ADI) was considered to be 0.003 mg/kg body weight while the acute reference dose (ARfD) was 0.03 mg/kg body weight (32).

## RESULTS AND DISCUSSION

### Quantification of levels of diazinon residues in the farm-gate baskets of *Brassica oleracea var. acephala* from the KOSFIP area of Homa Bay County for health risk assessment

Analysis of diazinon residues in the farm-gate *Brassica oleracea* var, *acephala* samples (Table 1) using the standard normal distribution function (f(z)) showed that 78% of all the samples had detectable levels of residual diazinon while 22% had non-detectable levels. Similarly, the percentage of farm gate samples with higher residues of diazinon than the acceptable 0.05 mg/kg was 70%. The findings were comparatively higher than levels reported by similar studies in Ghana (31), Nigeria (34) and other parts of Kenya (33,35), that reported trace levels of diazinon residues with less than 10% of the samples being above the Codex MRL.

**Table 1:**
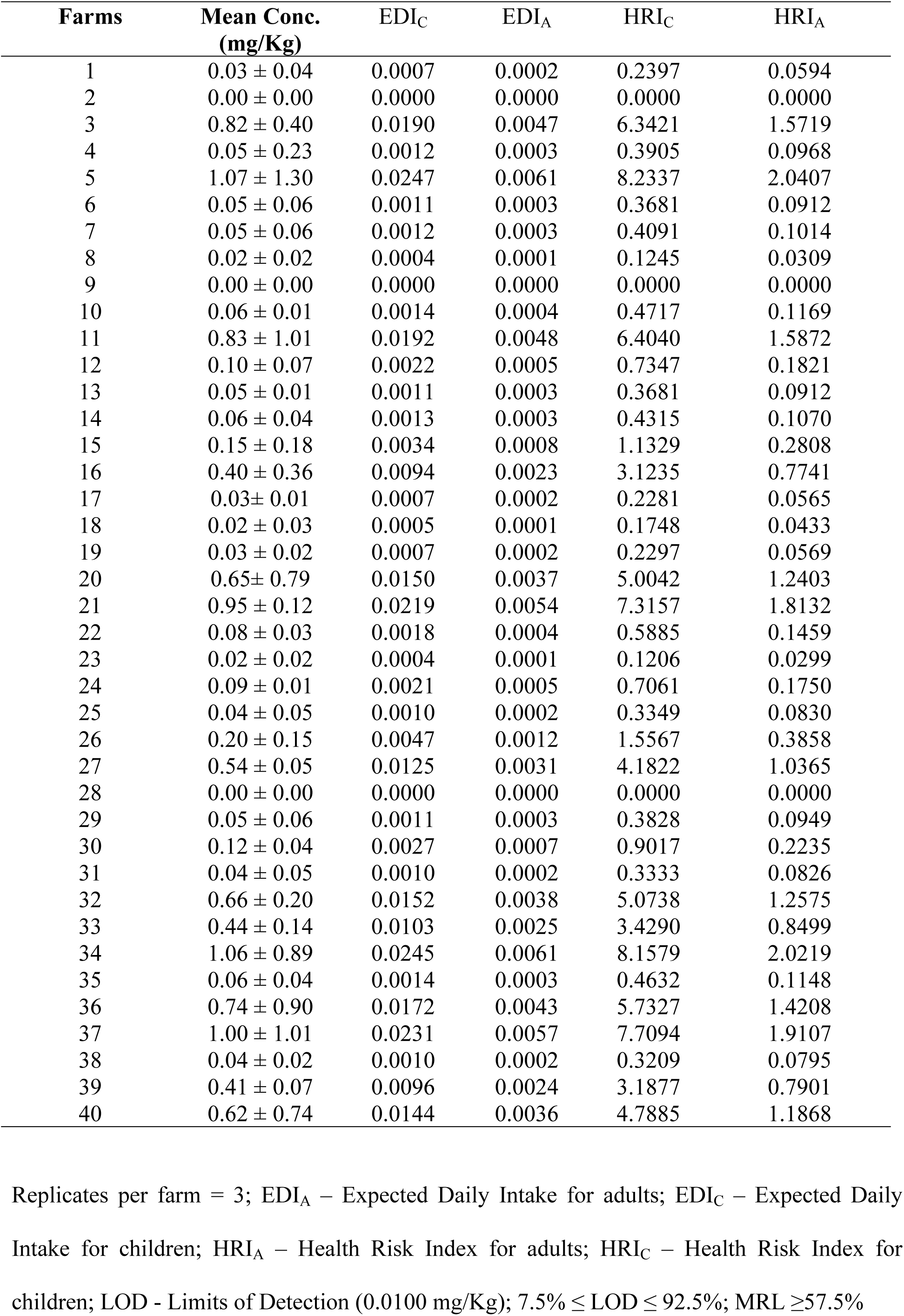
Levels of diazinon residues in farm-gate baskets of Brassica oleracea var. acephala from selected KOSFIP blocks and resultant EDI and HRI for children and adults.

The measures of central tendency (mean, median and mode) displayed a positively skewed distribution with a coefficient of variation (CV) of 122% (Table 2). The distribution demonstrated that the residues levels of diazinon in farm-gate *Brassica oleracea var. acephala* were highly variable with a large range. The variability could be as a result of multiple factors associated with inadequate training of farmers on good agricultural practices and poor surveillance by respective national authorities (38). Consequently, farmers in KOSFIP area require regular surveillance and training on use of diazinon. The findings also suggest that the recommended diazinon application conditions of rates and pre-harvest intervals may be too short and not suitable for the study area. In addition, the findings suggest that KOSFIP farm-gate vegetables need thorough washing before cooking to reduce the leaf surface residual levels on the vegetables. The data set (Table 1) had no outliers. Corresponding measures of central tendency and dispersion derived from the data set are shown in Table 2.

**Table 2:**
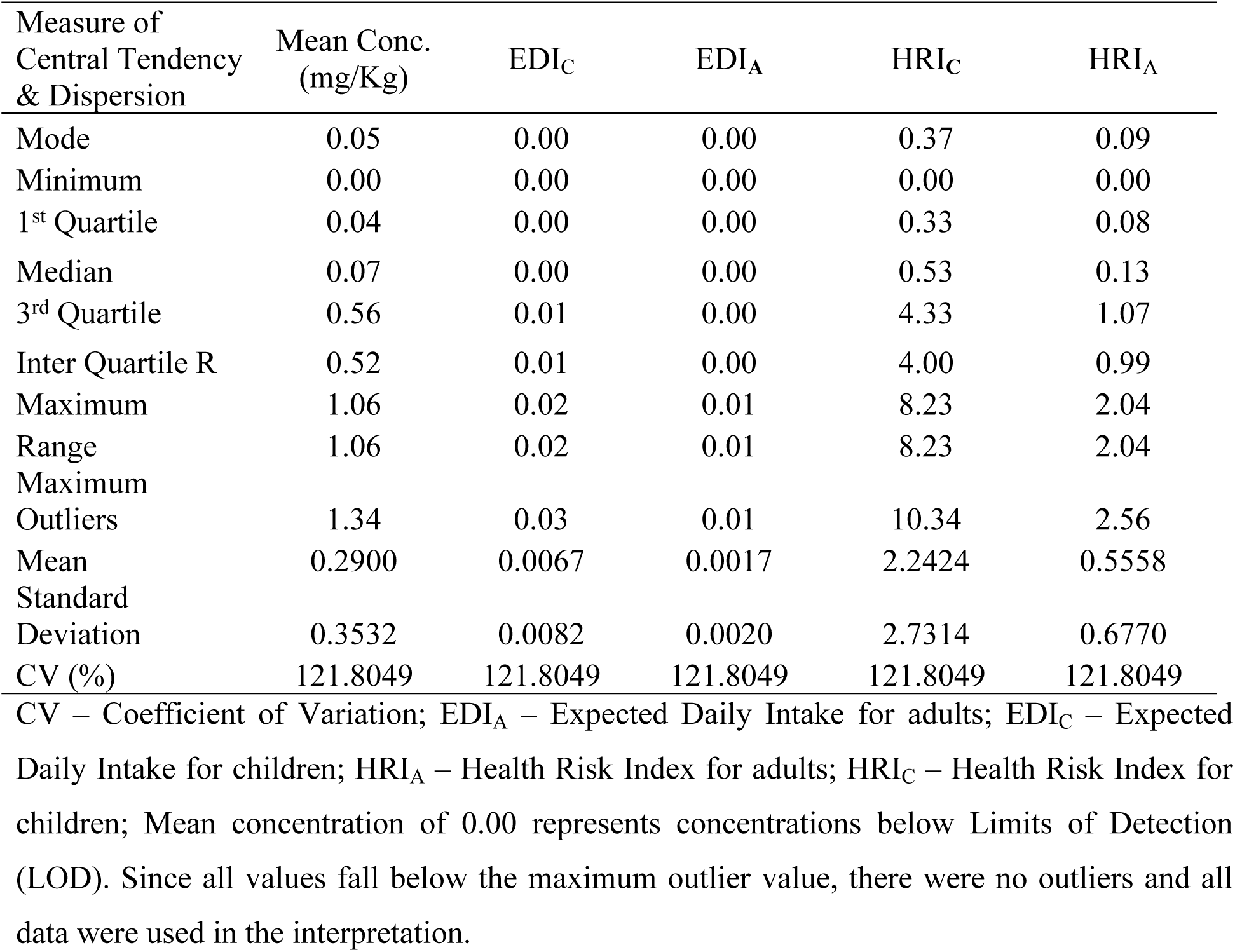
Measures of central tendency and dispersion for levels of diazinon residues in farm-gate baskets of Brassica oleracea var. acephala from KOSFIP area and resultant EDI and HRI for children and adults.

Given that food safety standards encourage infinitesimal residual levels of pesticide residues (17), a positively skewed distribution with all measures of dispersion falling below Codex MRL (0.05 mg/kg) would be most preferable. However, the findings of this study demonstrate mean residual diazinon concentrations much higher than Codex MRL. As a result, the high diazinon residues levels in the farm gate samples may pose health risks to consumers. Subsequently, most of the farm gate *Brassica oleracea var. acephala* vegetables treated with diazinon at KOSFIP may not therefore be safe for human consumption. The standard normal distribution function (f(z)) for the health risk indices showed that approximately 64% of the samples could pose health risks to children. Similarly, approximately 26% of the samples could pose health risks to adults. The findings indicate that children consuming *Brassica oleracea* var. *acephala* from the study area may be more at risk than adults. These findings were similar to the levels reported in some vegetables of Bangladesh (39), Kuwait (40), Nigeria (41), Ghana (42) and Sudan (43), where over 30% of samples reported residue levels above the Codex MRLs. The estimated daily intake (EDI_C_) and resulting health risk indices for children (HRI_C_) indicate that children consuming the vegetable from the study area have higher chances of developing diazinon related health problems (44,45). Given that data on residual levels with computed EDI and HRI for children are not available, comparisons of health risk factors for children in different regions have not been done.

On the other hand, the estimated daily intake for adults (EDI_A_) and resultant health risk index for adults (HRI_A_) were comparatively lower than ratios for children (Table 1 and Table 2). The findings were similar to residual levels of diazinon in cauliflower of Bangladesh (46), tomatoes of Spain (47) and Iran (48), respectively. The effect of the residual levels of diazinon on EDI and HRI for adults were higher than the findings in Chinese kale (49), spring onion, parsley onion and ginger vegetables in Thailand (50). In addition, the results were also higher than those reported for yard long bean in Bangladesh (46), apple in Pakistan (51), *T. occidentalis* and *C.argentea* in Nigeria (34), *Brassica oleracea var. acephala* (35,37) and tomatoes (Ngolo *et al*., 2019) in Kenya.

However, the results and the resultant EDI and HRI were lower than ratios reported on eggplant and tomatoes in Pakistan (39,52), watermelon (41), spinach and onions (53) in Nigeria, tomatoes in Ghana (33), cucumber and tomatoes in Sudan (43). Given the high variability displayed (coefficient of variation (C.V.) of 122%), the farmers and consumers of the vegetables are likely to be exposed to diazinon associated health risks (54). These health challenges may threaten human population in the study area. On the same note, though the adults have a lower mean ratio of 0.559, continuous consumption of this popular vegetable may cumulatively raise the exposure levels and result in devastating health impacts (55).

The presence of inappropriate levels of diazinon residues in the leaves of *Brassica oleracea var. acephala* may be a consequence of non-adherence to good agricultural practices (GAPs) such as failing to observe the application conditions of dosage and pre-harvest intervals. Such disregard are popular with smallholder farmers in developing countries for locally consumed vegetables (5,15,56) and in regions where surveillance and monitoring activities are inadequate. The unacceptable residue levels could be due to inability of the farmers to interpret the rates as prescribed on the labels or utter disregard of the rates, and inadequate surveillance of farmers on the use of pesticides. Inappropriate levels may also be attributed to inability of the farmers in the study area to consider other viable methods of pest control provided by Integrated Pest Management (IPM) guidelines (40,57) during heavy infestations. To manage the unacceptable residue levels and related health problems, training of farmers on GAP and alternatives to chemical pest control should be initiated in the study area. National regulatory agencies should equally strengthen surveillance on farmers with emphasis on observance of pre-harvest intervals and pesticide dosages. The general public should also be sensitized on the need to reduce the risks through proper culinary processing of the vegetables before consuming them. Lastly, the inappropriate residue levels may result from label application conditions which were extrapolated from field trials done in other regions but not suitable for the study area. It is therefore well justified to constantly monitor residual levels of diazinon in the vegetable and to develop pesticide safety level monitoring policies to mitigate resultant health problems due to unacceptable residue levels.

In conclusion, the findings in this study showed that most of the farm-gate *Brassica oleracea var. acephala* treated with diazinon within the KOSFIP area of Homa Bay County of Kenya had higher than tolerable residue levels. The residual diazinon quantities may pose significant health risks to the consumers. The high levels suggested that most farmers of *Brassica oleracea var. acephala* who use diazinon within the KOSFIP area may not be observing good agricultural practices (GAPs). Consequently, there is need to restrict use of diazinon on *Brassica oleracea var. acephala* at KOSFIP and to consider alternative pesticides with shorter PHIs to reduce high levels of residual diazinon in farm gate vegetables. In addition, there is need for a survey study into the extent of good agricultural practices (GAPs) with respect to pesticide use in the production of *Brassica oleracea var. acephala* and other vegetables within the KOSFIP area of Homa Bay County. On the same note, there is need for health risk index (HRI) determination for various age groups with estimated local weights, consumption rates and pesticide safety levels (PSL) adapted to the local environment. Finally, there is need for a comparative study on the effects of processing (washing, blanching, cooking) to establish if such processing methods significantly reduces diazinon residue levels in Brassica *oleracea var. acephala* of KOSFIP area.

This study has provided baseline information required for the establishment of Good Agricultural Practices (GAP) towards sustainable use of diazinon in production of *Brassica oleracea var. acephala* in the KOSFIP area. The study has provided a basis for discouraging the use of diazinon in the production of *Brassica oleracea var. acephala* in KOSFIP and other environments.

## Data Availability

All data produced in the present work are contained in the manuscript

## Acknowledgements

The authors appreciate Lillian Cherono of KOSFIP for her support as an agronomist, and Joab Oloo Onyango for provision of experimental plots within the KOSFIP scheme. In addition, the authors acknowledge the management of Kenya Plant Health Inspectorate Services (KEPHIS), and the KEPHIS Analytical Chemistry Laboratories technical team at Karen - Nairobi: Onesmus Mwniki, Lucy Nyamu, Samwel Maiyo, Edmond Momanyi, Aeline Kiptum, James Njenga, and Robert Njuguna Koigi. Finally, we appreciate the members of Prof Owuor’s research group and Dr. Charles Ochieng for their critique to the thought processes.

